# A Study on the Correlation between Adolescents’ Online Social Connections and Anxiety and Depression

**DOI:** 10.1101/2024.10.04.24314915

**Authors:** Li Mengxuan, Xu Jiajun, Liu Ying, Xu Jiaqi, Chen Hong

## Abstract

**OBJECTIVE:** To investigate the current status of anxiety and depression among adolescents and to analyze the correlation between adolescents’ online social connections and their depression and anxiety.

**METHODS:** The study adopted a cross-sectional survey research design, conducted in a middle school located in a city in Sichuan Province. Classes were selected through simple random sampling, with all students in the selected classes serving as study participants. Anxiety, depression, and online social connections were assessed using the Chinese version of the DSM-5 Anxiety Inventory for Children and Adolescents, the DSM-5 Depression Inventory for Children and Adolescents, and the Revised Social Connectedness Scale, respectively.

**RESULTS:** The prevalence rates of anxiety and depression among the participants were found to be 49.1% and 47.1%, respectively. A significant positive correlation was identified between online social connections and both anxiety and depression.

**CONCLUSION:** The findings indicate a concerning prevalence of anxiety and depression among adolescents. It is crucial for healthcare professionals to recognize the multifaceted factors contributing to these mental health challenges and to remain vigilant regarding adolescent mental health. Leveraging the impact of online social connections, early psychological intervention strategies should be implemented to promote healthy online relationships and mitigate the risks of anxiety and depression among adolescents.

## Preamble

Mental health challenges, particularly anxiety and depression, represent significant public health concerns among adolescents. Depression threatens adolescent mental health by increasing the risk of suicidal behavior not only during adolescence ^[1]^ but also serving as an indicator of potential suicidal tendencies in adulthood ^[2]^. In addition to this, anxiety is a key impediment to the overall development of adolescents, adversely affecting their physical health, academic performance, and social skills ^[3]^. Various factors contribute to anxiety and depression within this demographic, including gender, family income, parental education, parent-child relationships, study efficiency, and daily physical activity ^[4,5]^.

Social connectedness refers to the subjective sense of belonging and closeness experienced by individuals in their relationships with others and this feeling of connection can be shaped by their experiences of close relationships, such as with family, friends, and other social groups, as well as their experiences of estrangement from strangers or society as a whole ^[6]^. In today’s digital age, cybersocial bonding has emerged as a unique form of social connection. It refers to the extension of real social bonds into the realm of non-face-to-face interactions facilitated by electronic mediums, particularly online platforms. The National Research Report on Internet Use by Minors in 2021 revealed that an astonishing 96.8% of underage individuals in China are utilizing the Internet, indicating an extremely high level of internet penetration among this group ^[7]^. With the advent of the digital age, teenagers’ lives and studies have become inseparable from the online realm, and their social activities on the internet are increasing by the day. Overseas studies have shown a correlation between online social connections and mental health issues such as anxiety and depression among adolescents ^[8,9]^. However, the findings regarding this correlation have been inconsistent, with varying degrees of strength. Therefore, this study aims to investigate adolescents in order to gain insight into the relationship between online social connections and anxiety and depression. The results of this study will provide a scientific basis for implementing targeted interventions to promote the physical and mental well-being of adolescents.

## 1. Objects and Methods

### 1.1 Subject of the Study

This study is a cross-sectional survey research conducted in Bazhong City, Sichuan Province, in September 1, 2022 - October 31, 2022. A middle school was selected using a simple random sampling method, and nine classes were chosen to represent the sample population. All eligible students in the selected classes were invited to participate in the survey. Prior to the survey, the purpose, methodology, and significance of the study were explained to the respondents. To ensure full understanding, both the respondents and their guardians were informed of the specific content and purpose of the survey. Written informed consent has been obtained from the guardians. Formal ethical approval was obtained from the Ethics Committee of West China Hospital, Sichuan University (Approval No. 2019-907). According to the sample content estimation method proposed by Kendall, the sample size should be 10-20 times the number of independent variables. Considering that this study involves 22 independent variables, a sample size of 220-440 was determined. A total of 402 questionnaires were distributed, and 397 valid questionnaires were returned, resulting in an effective recovery rate of 98.8%.

The inclusion criteria for this study were as follows: (1) age between 11 and 17 years old, and (2) willingness to participate in the psychological assessment survey, with a signed informed consent form from their guardians.

The exclusion criteria were as follows: (1) individuals with severe physical illnesses, (2) individuals with severe mental disorders, (3) individuals with intellectual developmental disorders, and (4) individuals with hereditary diseases or peripheral nerve disorders.

### 1.2 Research Tools

#### 1.2.1 General Information Collection Questionnaire

The general information collection questionnaire was designed by the researcher to gather demographic data from the study participants. Information such as gender, age, place of residence, only child status, previous illnesses, current medications, and family financial situation were included. The content of the questionnaire was reviewed and revised by experts on the research team.

#### 1.2.2 Social Connection Scale Revised (SCS-R)

The Social Connection Scale Revised (SCS-R) was originally developed and refined by Lee et al.^[10]^. Fan Xiaolan et al.^[11]^ then translated and further revised the scale. It consists of 20 items, divided into two main sections: social connectedness and social disconnectedness. In the section on social connectedness, items 1, 2, 4, 5, 8, 10, 12, 14, 16, and 19 are included. In the section on social disconnectedness, items 3, 6, 7, 9, 11, 13, 15, 17, 18, and 20 are included, with reverse scoring applied to all the items in this section. The scale utilizes a six-point Likert scale, where each item is scored from “1” to “6”. A score of “1” indicates “Strongly Disagree” and a score of “6” indicates “Strongly Agree”. Higher scores indicate a greater perceived closeness in interpersonal situations. The Cronbach’s alpha coefficient for the SCS-R was calculated to assess the internal consistency of the scale. The coefficient was found to be 0.911 for the total social connectedness scale, 0.822 for the social connectedness dimension, and 0.867 for the social disconnectedness dimension.

#### 1.2.3 DSM-5 Anxiety Scale for Children and Adolescents Chinese Version

The DSM-5 Anxiety Scale for Children and Adolescents is specifically designed for assessing generalized anxiety symptoms in adolescents aged 11 to 17 years. The English version of the scale has obtained official licensure from the Patient Measurement Reporting Information System Health Organization (PROMIS Health Organization or PHO). To cater to Chinese-speaking populations, the Personality and Psychopathology Research Group from the School of Psychology and Cognitive Science at Peking University undertook the translation of the scale. Subsequently, Zhang Yewen et al ^[12]^ revised and enhanced the content of this Chinese version. The scale comprises 13 items, each aiming to evaluate the extent of anxiety experienced by the adolescent over the past week. Employing a five-point Likert rating system, respondents rate the frequency of occurrence for each item, ranging from 1 (never) to 5 (almost always). The total score of the scale ranges from 13 to 65, whereby scores equal to or exceeding 28 are indicative of the presence of anxiety symptoms. Moreover, a higher score indicates a greater severity of anxiety experienced by the individual. The internal consistency of the scale, as measured by Cronbach’s alpha coefficient, was found to be 0.90 for the total scale. The Cronbach’s alpha coefficients for the general anxiety and situational anxiety factors were 0.89 and 0.73, respectively. The scale also demonstrated good test-retest reliability, with a coefficient of 0.78 for the total scale. Additionally, the test-retest correlation coefficients for the two factors were 0.63 and 0.67, respectively.

#### 1.2.4 DSM-5 Depression Scale for Children and Adolescents

The DSM-5 Depression Scale for Children and Adolescents is a recognized instrument designed to evaluate the severity of depressive symptoms in individuals aged 11 to 17. The scale contains 14 different items, each of which has 5 rating levels: 1 for “never”, 2 for “almost never”, 3 for “sometimes”, 4 for “often”, and 5 for “Almost Always”. Total scores on the scale range from 14 to 70, with a score of ⩾32 indicating the presence of depressive symptoms. Higher scores on the scale correspond to more severe levels of depression. The reliability coefficient for internal consistency, as measured by Cronbach’s alpha, was found to be 0.95 in this particular study. Additionally, the KMO measure of sampling adequacy yielded a value of 0.947, indicating high suitability of the scale for assessment purposes.

### 1.3 Quality Control

Researchers undergo standardized training and adhere to uniform instructions during the research process. The questionnaires are completed in a controlled environment, with measures in place to protect the confidentiality of the responses. To facilitate efficient data entry, a comprehensive data management system is developed. This system incorporates quality control measures, such as predefined value ranges and logical checks, to ensure data accuracy and reliability. Moreover, a robust data cleansing and validation procedure is established, performed by highly skilled personnel, to guarantee the consistency of data management and analysis.

### 1.4 Statistical Analysis

The data entry process was conducted utilizing the Excel software, while the analysis of the collected data was performed using SPSS 26.0. The descriptive analysis of count data involved presenting the frequency counts and constitutive ratios. Furthermore, to examine the relationship between online social connections and anxiety among adolescents, Spearman’s correlation analysis was employed.

## 2. Results

### 2.1 Analysis of General and Demographic Information

A total of 402 questionnaires were collected. After excluding 5 invalid questionnaires, 397 valid questionnaires were obtained, resulting in an effective recovery rate of 98.8%. Of the respondents, 230 were male and 167 were female, resulting in a sex ratio of 1.38. The mean age of the participants was (13.95±1.01) years. Please refer to Table 1 for further details.

**Table 1.**
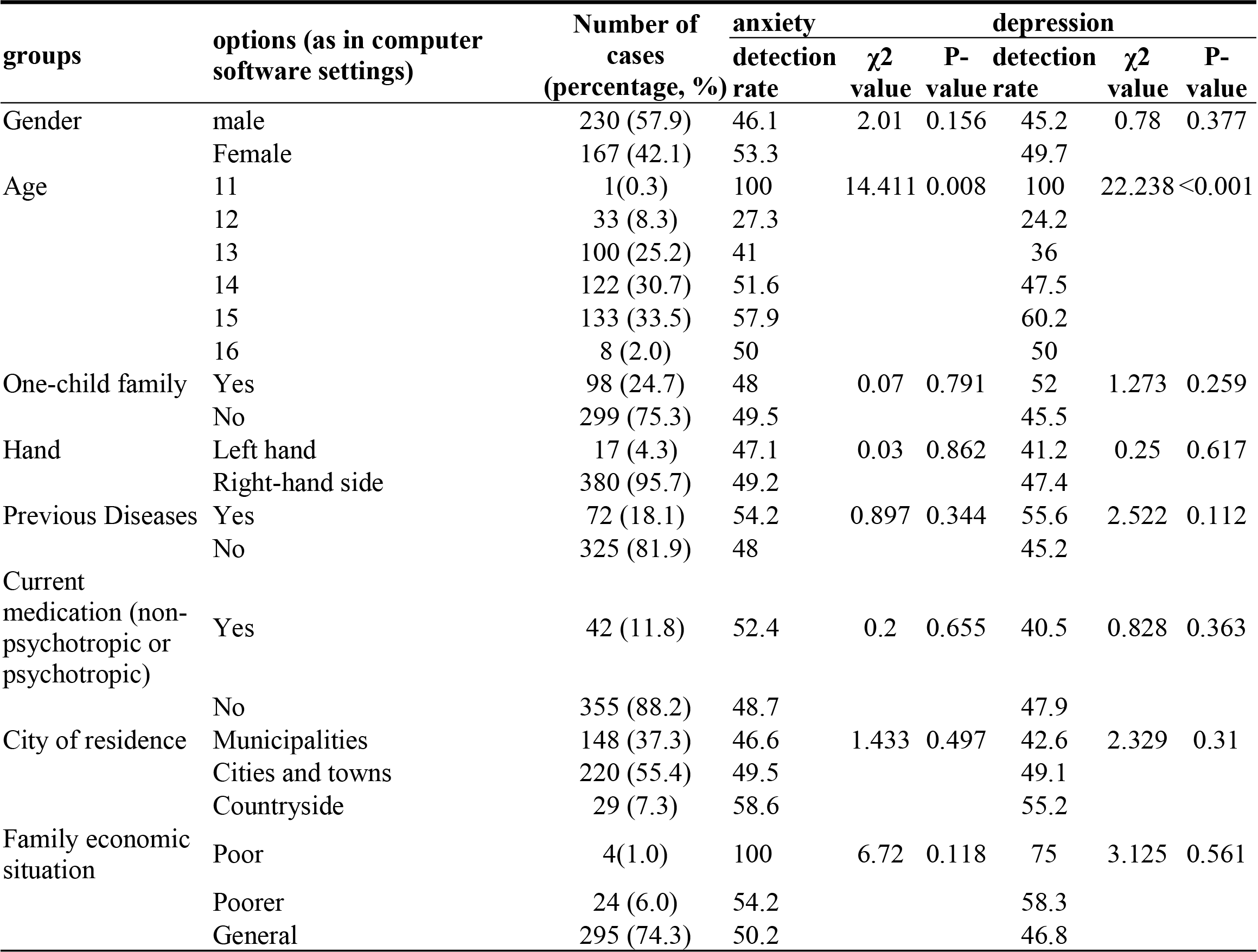
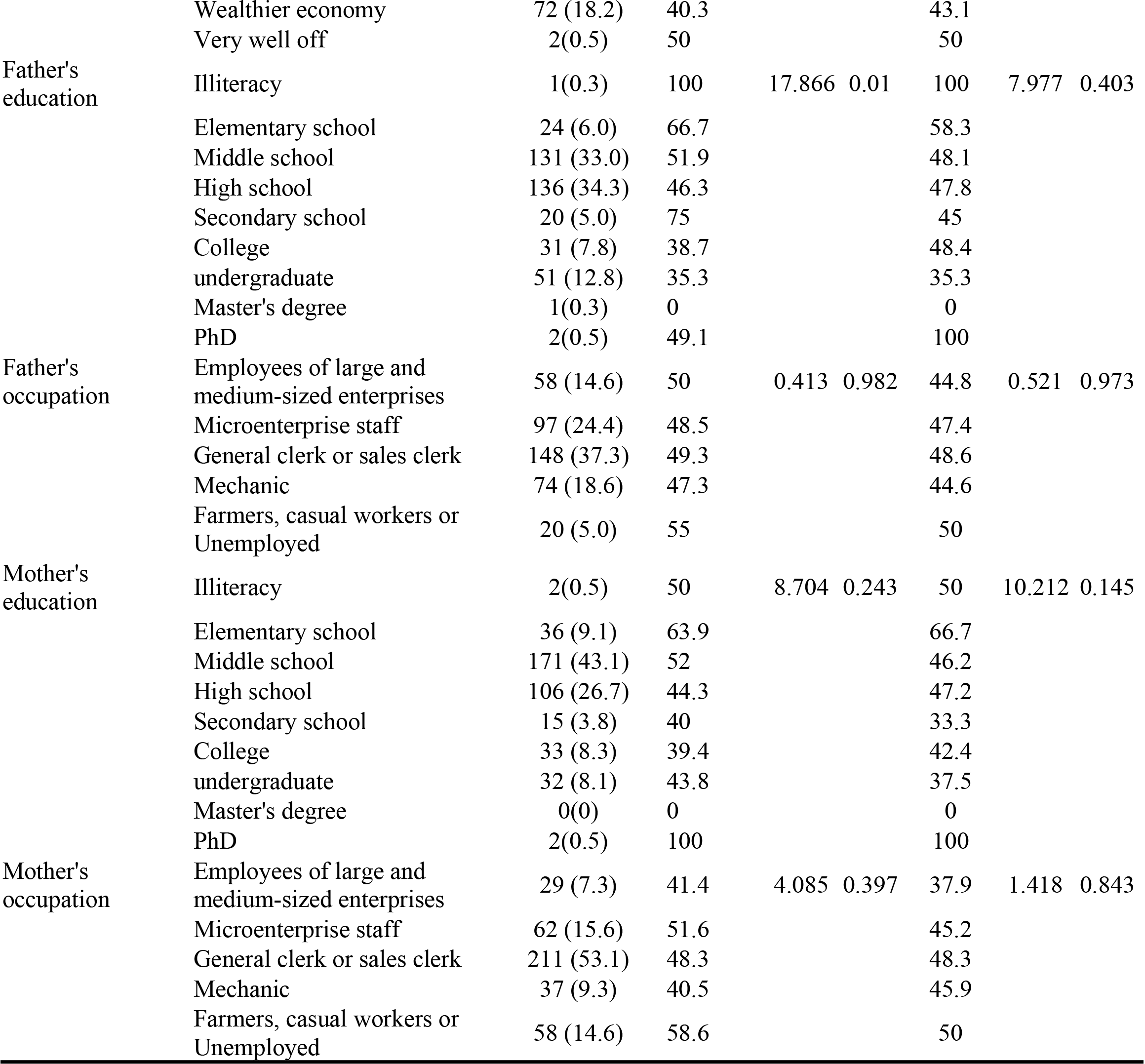
Comparison of Anxiety and Depression Detection Rates in Different Subgroups of Adolescents.

### 2.2 Detection Rate of Anxiety and Depression

Among the adolescents surveyed, 195 individuals were detected with anxiety symptoms, resulting in a detection rate of 49.1%. Additionally, 187 adolescents were detected with depressive symptoms, resulting in a detection rate of 47.1%. Please refer to Table 2 for further details.

**Table 2.** The number and detection rate of anxiety and depression in adolescents.

|  | aggregate | number of<br>people<br>suffering<br>from a<br>disease | detection<br>rate |
| --- | --- | --- | --- |
| anxiety | 397 | 195 | 49.1 |
| depression | 397 | 187 | 47.1 |

### 2.3 Correlations between Anxiety, Depression, and Online Social Connections

The results presented in Table 3 demonstrate a significant and positive correlation between online social connections and anxiety. Similarly, network social connectedness is significantly and positively correlated with depression.

**Table 3.** Analysis of the correlation between adolescents’ online social connections and anxiety and depression.

|  | online social connections | anxiety | depression |
| --- | --- | --- | --- |
| online social connections | 1.000 |  |  |
| anxiety | 0.338** | 1.000 |  |
| depression | 0.423** | 0.733** | 1.000 |

### 2.4 Analysis of Factors Influencing Anxiety and Depression in Adolescents

In order to examine the factors that influence anxiety and depression in adolescents, stratified regression analyses were performed. The dependent variables were anxiety and depression, and the independent variables included in the analysis were determined based on their statistical significance (p<0.05). The impact of online social connections on adolescent anxiety and depression was assessed, and the results indicated that this factor independently accounted for 13.3% of the variance in adolescent anxiety and 19.7% of the variance in depression. Detailed findings can be found in Tables 4 and 5.

**Table 4.**
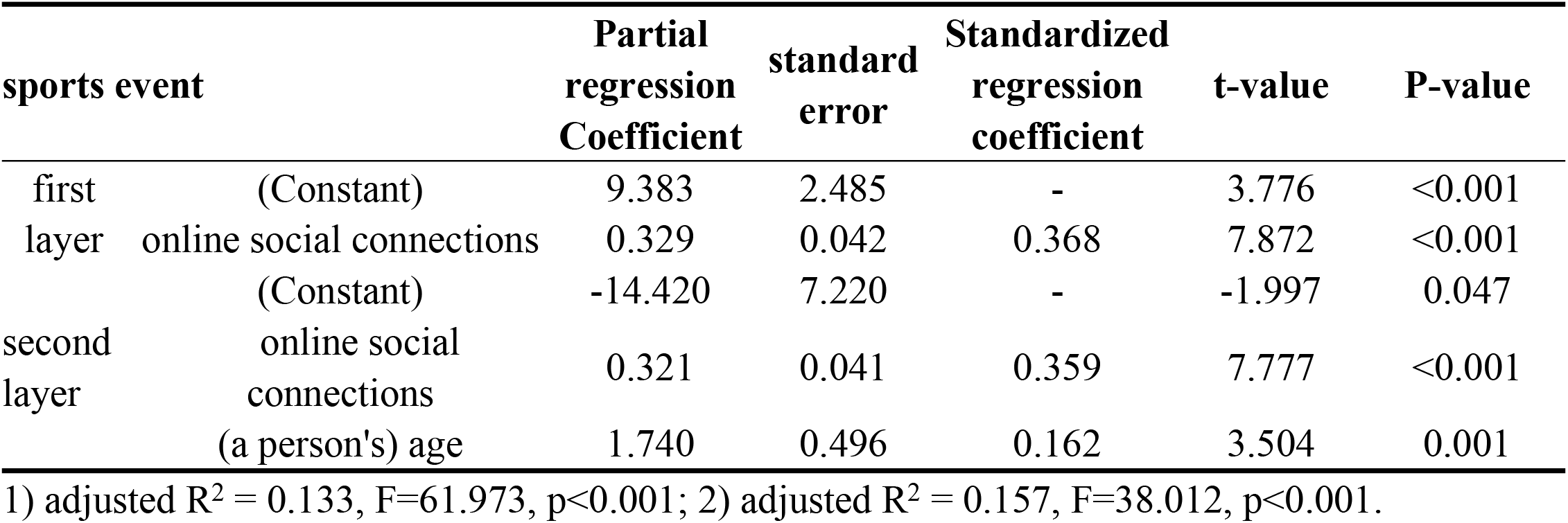
Stratified regression analysis of factors influencing factors of adolescent anxiety.

**Table 5.**
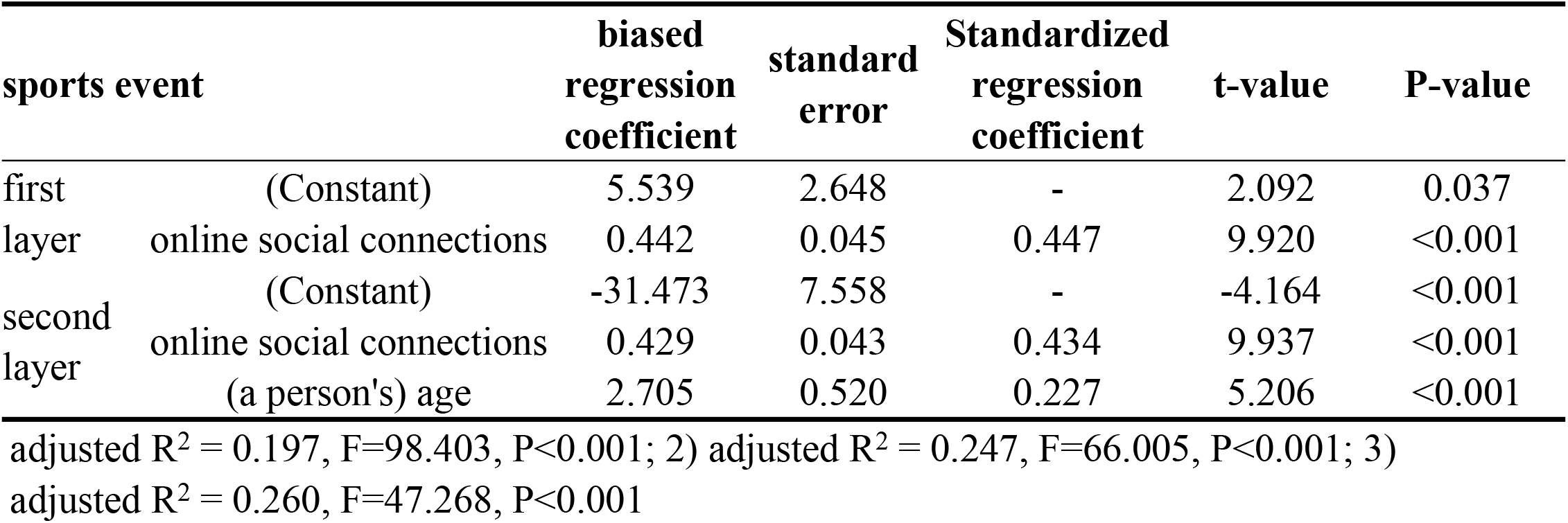
Stratified regression analysis of factors influencing factors of adolescent depression.

| sports event |  | biased regression coefficient | standard error | Standardized regression coefficient | t-value | P-value |
| --- | --- | --- | --- | --- | --- | --- |
| first | (Constant) | 5.539 | 2.648 | - | 2.092 | 0.037 |
| layer | online social connections | 0.442 | 0.045 | 0.447 | 9.920 | <0.001 |
|  | (Constant) | -31.473 | 7.558 | - | -4.164 | <0.001 |
| second | online social connections | 0.429 | 0.043 | 0.434 | 9.937 | <0.001 |
| layer | (a person's) age | 2.705 | 0.520 | 0.227 | 5.206 | <0.001 |

adjusted $R^2 = 0.197$ , $F=98.403$ , $P<0.001$ ; 2) adjusted $R^2 = 0.247$ , $F=66.005$ , $P<0.001$ ; 3)
adjusted $R^2 = 0.260$ , $F=47.268$ , $P<0.001$

## 3. Discussion

### 3.1 Anxiety and Depression Levels in Adolescents

The present study yielded noteworthy findings regarding the detection rates of anxiety and depression in adolescents. The prevalence of anxiety detection was found to be 49.1%, whereas the prevalence of depression detection was 47.1%. A comprehensive meta-analysis encompassing data from 27 countries reported that the prevalence of anxiety among secondary school students was 6.5%, whereas the prevalence of depression was 2.6% ^[14]^. Additionally, a study conducted in China by Liu Xiaoqun et al.^[15]^ revealed detection rates of 15.1% for anxiety and 20.4% for depression. Another study conducted in Hunan Province, involving 10,220 secondary school students, indicated detection rates of 11.7% for anxiety symptoms and 15.3% for depressive symptoms over a two-week period. The detection rates of anxiety and depressive mood observed in our study were comparatively higher than those reported in the aforementioned studies. One possible explanation for this discrepancy could be attributed to the utilization of the DSM-5 Anxiety and Depression Scale for Children and Adolescents, which is renowned for its heightened sensitivity. Moreover, it is important to acknowledge that adolescents traverse an intricate phase characterized by substantial academic pressures, coping with physical development and peer interactions, rendering them more susceptible to experiencing anxiety and depression. Furthermore, the unprecedented challenges posed by the COVID-19 pandemic exacerbate the psychological strain encountered by adolescents. The implementation of prolonged online learning has deprived them of the familiar school and classroom environment, consequently reducing their opportunities for peer interaction. Consequently, this sense of isolation and uncertainty regarding knowledge acquisition may facilitate the development of depression and anxiety among adolescents.

### 3.2 Anxiety is Positively Associated with Online Social Connections

The findings of this study reveal a positive relationship between adolescent anxiety and online social connections. Individuals with anxiety tend to compensate for the lack of offline relationships by actively engaging in their social lives within the online sphere ^[16]^. Moreover, research has demonstrated that for individuals with high levels of anxiety, the correlation between online social interactions and emotional support, as well as the correlation between online social interactions and the desire to socialize, is much stronger ^[17]^. Anxious individuals may increase their online interactions as a means of avoiding face-to-face interactions ^[18]^. The online environment provides a sense of comfort and serves as a primary social channel, often replacing face-to-face interactions.

### 3.3 Depression is Positively Correlated with Online Social Connections

The results of the correlation analysis of this study show that adolescent depression is positively associated with online social connection. The study show that depression tends to be more prevalent among individuals who engage in prolonged online media use ^[9]^. However Nakagomi et al ^[19]^ have argued that using the internet for communication actually has a protective effect on the likelihood of developing clinical depression. It is worth noting, though, that higher levels of depressive symptoms have been found to predict a smaller number of friends on Facebook and decreased contact between friends, albeit to a lesser extent on average ^[20]^. These inconsistencies may be attributed to the fact that excessive immersion in the online environment reduces the time spent engaged in face-to-face interactions and limits opportunities for social engagements, thereby impacting the risk of depression.

## 4. Limitations

There are several limitations associated with this study. Firstly, the generalizability or representativeness of the findings may be somewhat constrained due to the exclusivity of the sample, which was obtained solely from a single educational institution. To enhance the validity and applicability of the results, it would be beneficial to increase the sample size in future studies and extend the research to encompass a broader range of schools and regions. Secondly, in order to obtain a more comprehensive and profound insight into the study subject, it is imperative to validate the findings by replicating the research in different geographical locations. This expansion would allow for a broader examination of the phenomenon, leading to a more robust and nuanced understanding.

## 5. Conclusion

The findings of this study indicate a high prevalence of anxiety and depression symptoms in adolescents, with a noteworthy positive association between online social connections and anxiety and depression. Prolonged engagement in online interactions among adolescents may result in tangible social isolation, consequently elevating the risk of anxiety and depression. Thus, it is necessary for practice to evaluate the internet usage patterns and mental well-being of adolescents in order to identify any existing issues. Through educational initiatives and training programs, we can aid adolescents in cultivating healthy online habits and prevent mental health complications. For those adolescents who have already encountered mental health problems, we can offer psychological support and guide them towards professional assistance, thereby facilitating the development of effective interventions.

## Data Availability

All relevant data are within the manuscript and its Supporting Information files.

